# Are automated documentation-error judges fit to measure ambient AI scribes? A pre-registered, blinded human-validation study

**DOI:** 10.64898/2026.08.14.26360441

**Authors:** Henry Bergman, Vivian Liu, Ben Austin, Rohan Sangera

**Affiliations:** Clinical AI Research, Heidi Health, 140 Goswell Road, London EC1V 7DY, United Kingdom

## Abstract

**Objectives:** Safety claims for ambient artificial intelligence (AI) scribes rest on automated judges that detect documentation errors and grade clinical risk. Expert reviewers are under-sensitive and disagree with one another, so no gold standard exists and validation cannot mean accuracy. We tested whether such judges are a defensible instrument: reproducible, within the envelope of expert disagreement, and non-differential across arms.

**Methods:** Pre-registered, blinded validation study nested in a multi-country simulation of ambient AI documentation (English setting), reported per GRRAS. Ten external clinicians independently adjudicated a stratified sample of 434 pipeline flags, retained and screen-discarded, blinded to note authorship, identification source, the pipeline’s verdict and severity tier. Agreement used Gwet’s AC1; proportions carry Wilson intervals. Three propositions were pre-specified: envelope parity, non-differential behaviour across arms, and concordance on consensus cases.

**Results:** All ten reviewers completed: 565 adjudications across 434 items, 131 of them double-rated. Inter-clinician agreement on genuineness was fair (raw 59%, 95% CI 50 to 67; AC1 0.24), leaving no human consensus to serve as truth. Judge-clinician agreement was 64% (95% CI 60 to 68), overlapping that interval. Behaviour was near-symmetric on contrast-critical metrics: kept-precision 74% for AI against 81% for clinician notes, and severity signed gap +0.06 against -0.09 tiers. One sub-metric was asymmetric: removed-confirmed 56% against 42%, so the screen over-removes more on clinician notes, a direction conservative to the parent contrast. On 77 consensus items the pipeline concurred on 70% (95% CI 59 to 79). Latent-class triangulation placed the genuine-error rate among flagged candidates at 68% (94% credible interval 48 to 83).

**Conclusions:** The judges behave as a consistent, near-non-differential, clinician-equivalent instrument. This licenses a directional AI-versus-clinician contrast under a non-differential misclassification argument, subject to its conditions. It is not a claim of accuracy, which moderate consensus concordance and fair reliability preclude, and the genuine-error rate is best reported as an interval.

**What is already known on this topic:**

- Ambient AI scribes are entering routine practice, and their evaluation depends on detecting documentation errors at a scale that unaided human review cannot achieve
- Large language models have been validated as judges of clinical text quality, with strong agreement against human raters on ordinal quality instruments
- Human record review is known to be under-sensitive, and expert reviewers agree only moderately even on explicit quality criteria, so no gold standard for documentation error exists

**What this study adds:**

- Clinicians agreed only fairly on whether a flagged item was a genuine documentation error (AC1 0.24), which is substantially weaker than published agreement on ordinal note-quality scoring and indicates that error detection is the harder judgement.
- An automated judge agreed with clinicians about as well as clinicians agreed with one another, and behaved near-symmetrically across AI-authored and clinician-authored notes on the metrics that determine a comparative contrast.
- The one asymmetry detected runs against the sponsor’s interest, and the genuine-error rate among flagged candidates is identifiable only as an interval.

**How this study might affect research, practice or policy:**

- Where no gold standard exists, validation of automated documentation-error judges should be framed as defensibility, meaning reproducibility, parity with the expert envelope and non-differential behaviour, in place of accuracy.
- Comparative studies of AI and clinician documentation should report the direction in which any measurement asymmetry biases their own contrast.
- Absolute documentation-error rates derived from automated judges should be reported as bounded intervals.

## Introduction

Ambient AI scribes now draft clinical documentation directly from the consultation, and are being adopted at scale with early trial evidence of reduced documentation time.^1 2^ Their safety turns on a single empirical question: how often, and how seriously, does the generated note depart from what actually happened, either by asserting something that was not said or by omitting something that was. Comparative evaluations report both classes of error at non-trivial rates: a blinded specialty-reviewer study of 97 encounters detected hallucinations in 31% of ambient notes and 20% of physician-authored notes,^3^ and dedicated frameworks now exist for quantifying hallucination and omission in generated clinical summaries.^4^ Quantifying them at scale is a prerequisite for any claim that such systems are safe to deploy, and for any meaningful comparison with the documentation clinicians produce unaided.

That quantification has a measurement problem at its core: detecting a documentation error requires a reviewer, and every available reviewer is imperfect. Human expert review is the conventional reference, but it is expensive, slow and attention-limited. A clinician reading a fluent note misses much of what a line-by-line comparison against the transcript would surface, and this under-sensitivity is well documented in patient-safety measurement more broadly, where systematic detection identifies roughly ten times the adverse events found by conventional review.^5^ Reviewers also disagree with one another. A systematic review of 26 case-note audit studies found mean kappa values between 0.32 and 0.70, with reliability higher where criteria were explicit and where the judgement concerned outcome, and lower where criteria were implicit and the judgement concerned process error.^6^ Documentation-error adjudication sits at the harder end of both axes. Under-sensitivity is compounded by automation bias, whereby reviewers accept plausible machine output and detect fewer errors within it.^7^ Analyses of speech-recognition documentation show the same pattern, with only a minority of errors clinically significant and most corrected before signature.^8^

Large language model judges can compare note to transcript exhaustively and reproducibly, and have been validated against human raters for ordinal clinical-text quality scoring with strong inter-rater reliability.^9^ They are nonetheless suspected of the opposite failure to human reviewers: over-flagging, and in particular penalising the routine clinical concision that makes a note usable. Overlap-based automatic metrics are known to be insufficient proxies for clinical text quality,^10^ and model outputs are themselves stochastic, so identical inputs can yield different judgements.^11^ Neither instrument is ground truth, and, as we show, expert reviewers do not agree with one another, so there is no human consensus to appoint as truth in their place. The genuine error rate therefore lies somewhere between the under-sensitive human estimate and the over-sensitive automated one, and that location is not point-identifiable from the data.

The parent study on which this validation is nested is a multi-country simulation in which standardised actors and clinicians generated matched AI-authored and clinician-authored notes from the same simulated consultations across five language settings. Its error-rate and clinical-risk estimates are produced by a three-stage automated pipeline: identification of candidate errors; a false-positive screen that trims over-flagged candidates to genuine documentation errors; and a severity-grading panel that assigns each surviving error a clinical-risk tier. Every safety claim in that study rests on these judges behaving reasonably. The judges are the sponsor’s own, and industry-sponsored studies more often report favourable results and conclusions in a way conventional risk-of-bias appraisal does not capture,^12^ so the validation has to be designed against that hazard.

Because no gold standard exists, validation here cannot mean demonstrating that the judges are accurate. It means establishing that they are a defensible measurement instrument: reproducible, operating within the envelope of legitimate expert disagreement, and, most importantly for a study whose headline is a contrast between AI and clinician notes, non-differential, meaning not biased toward one arm of that contrast. We therefore frame validation as a falsifiable test of three propositions: that the judges (A) agree with clinicians about as well as clinicians agree with each other; (B) behave the same way on AI-authored and clinician-authored material; and (C) concur with clinicians on the cases where clinicians themselves concur. Failure of (B) or (C) would disprove fitness for purpose.

## Methods

This study is reported in accordance with the Guidelines for Reporting Reliability and Agreement Studies (GRRAS),^13^ which is the primary applicable standard, with DECIDE-AI followed for description of the system under evaluation and for assessor independence.^14^ No reporting guideline currently addresses the validation of an automated judge used as a measurement instrument, and we note that gap explicitly.

### Design and relationship to the parent study

This is a blinded, independent human-adjudication study nested within the parent simulation, which is its sole data source. All items adjudicated here are candidate errors produced by the parent study’s pipeline on simulated consultations; no new consultations were generated. Validation was restricted to the English setting to avoid confounding instrument behaviour with cross-language transcription artefacts, and because the parent study’s per-language error profiles indicated this setting as representative for a first-pass instrument validation.

### The instrument under validation

The pipeline operates in three stages, each pinned to fixed model versions and prompts for reproducibility. In identification, two parallel independent arms generate candidate errors: a human-identification arm, comprising flags raised by clinicians performing the parent study’s documentation-review task and retained where supported on re-checking, and an automated note-versus-transcript judge tuned to favour sensitivity over precision. In false-positive reduction, a single shared screen is applied to both arms, classifying each candidate as a genuine documentation error or a defensible non-error, meaning a faithful paraphrase, a reasonable inference, or appropriate clinical concision; the same screen is applied identically to AI-authored and clinician-authored notes. In severity grading, surviving errors are graded by a three-model panel on a Severity x Likelihood scale aligned to medical-device risk-management practice,^15 16^ with the tier taken from panel consensus. This study validates the false-positive screen and the severity panel, the two stages that determine the parent study’s reported error rates and risk tiers.

### Sample

We drew a stratified, deterministic sample of 434 individual candidate flags, designed so that every quantity of interest is estimable. Block A comprised 236 retained flags that the pipeline judged genuine, sampled across severity tiers and censusing the scarce Critical and High errors, to support both the post-screen precision estimate and the severity comparison. Block B comprised 198 flags the screen discarded, stratified by identification source and error type, to estimate the screen’s cost in genuine errors wrongly removed and its yield. Across both blocks the sample is stratified over identification source, note arm and error type. Thirty per cent of items (n=131) were independently rated by two reviewers to estimate inter-rater reliability, with double-rating stratified across type, tier, source and keep-or-remove decision. Sample size was fixed by the precision required on the expert envelope: 131 double-rated items give a 95% Wilson half-width of approximately 8 percentage points on raw inter-clinician agreement, which is sufficient to locate the envelope and leaves the AC1 point estimate correspondingly imprecise.^17^

### Reviewers, blinding and presentation

Ten external clinicians, all post-certification general practitioners with emergency and surgical representation, independently adjudicated the sample. Reviewers were blinded to note authorship, to the identification source, to the pipeline’s verdict on each item and to the severity tier, and each item was judged on its own evidence. A calibration step on shared worked examples preceded adjudication to limit between-reviewer drift. Items were presented one at a time in a self-contained browser instrument showing the flagged content, a neutral one-line consultation context, the verbatim excerpts highlighted in note and transcript, on-demand access to both in full, and the scoring rubric. To force a usable signal the genuineness decision was binary, with no uncertain option, and severity and likelihood were elicited only for items judged genuine.

### Reference standard

We treat the clinician panel as an imperfect comparator, without appointing it ground truth. This is a deliberate methodological position: expert reviewers are under-sensitive and, as our inter-rater analysis quantifies, agree with one another only weakly, so no human-derived truth is available against which to compute conventional accuracy. All estimands are therefore framed as characterisations of instrument behaviour relative to the expert envelope, or as bounds on quantities that are not point-identifiable.

### Risk-grading rubric

Risk was scored as Severity x Likelihood, each on a 1 to 5 scale giving a product of 1 to 25, with tiers Critical 16 to 25, High 10 to 15, Moderate 5 to 9 and Low 1 to 4, on the same rubric used by the parent study’s panel^15 16^. Severity captures the potential clinical consequence were the error to reach the record and influence care. Likelihood is operationalised as the probability that the error survives the workflow’s safety nets, including clinician editing before filing, detection on reading, and actionability, and goes on to affect care. The two axes were elicited and analysed separately, as likelihood is the more workflow-contingent and the less reliable of the two.

### Estimands, analysis and decision rule

Analyses are organised around the three propositions. Binary agreement uses Gwet’s AC1, which is prevalence-robust and avoids the paradoxical deflation of kappa at the skewed prevalences expected here.^18 19^ That prevalence dependence is also what makes raw and chance-corrected agreement diverge in case-note audit,^6 17^ so both are reported. Ordinal severity agreement was pre-specified as weighted kappa^20^ with the mean signed difference, the latter chosen to expose directional bias that a symmetric coefficient would conceal. Proportions carry Wilson confidence intervals.^21^ Agreement coefficients are interpreted against conventional benchmarks.^22^

For proposition A, clinician-clinician agreement on the double-rated items establishes the width of the expert envelope, against which judge-clinician agreement is compared. Because expert agreement is expected to be low, parity is necessary context, which falls short of proof of quality. For proposition B, we compare the screen’s human-confirmed precision and its removal-confirmation rate between arms, and the panel-versus-clinician signed tier difference between arms. For proposition C, we restrict to items on which a clinician supermajority agrees, where a local reference exists, and test whether the judge concurs; this is the only analysis capable of accuracy-like evidence, and the consensus subset may be small.

The pre-specified decision rule was that the judges are reasonable to use if they are non-differential across arms on both detection and severity, and concur with clinician consensus within the consensus clinicians’ own agreement band, with parity to the expert envelope reported as context.

They are deemed not fit for purpose if behaviour i s differential by arm, or if the judge systematically diverges from clinician consensus. A positive result supports the judges as a consistent, non-differential, defensible instrument, and explicitly does not establish accuracy.

### Pre-registration and reproducibility

The analysis plan, sample definition and seed, rubric and decision rule were fixed before data collection. The instrument, stratified sampler and analysis code are deterministic and archived, and the reviewer-facing material and item-level key are retained to permit exact reconstruction.

## Results

### Response

All ten invited reviewers completed their assignments, yielding 565 adjudications across 434 items, with 131 items independently double-rated.

### No gold standard: the judge sits within the expert envelope

Expert reviewers agreed only fairly on whether a flagged item was a genuine error: raw agreement 59% (95% CI 50 to 67) and Gwet’s AC1 0.24 on 131 double-rated items, which falls in the fair band on conventional benchmarks.^22^ There is therefore no human consensus that can serve as ground truth. The automated judge’s agreement with clinicians was 64% (95% CI 60 to 68), an interval that overlaps the inter-clinician interval throughout its upper half, so the judge behaves like one more reviewer drawn from the same panel (figure 1). The two intervals overlap widely enough that this comparison remains descriptive, falling short of a demonstration of equivalence.

**Figure 1.**
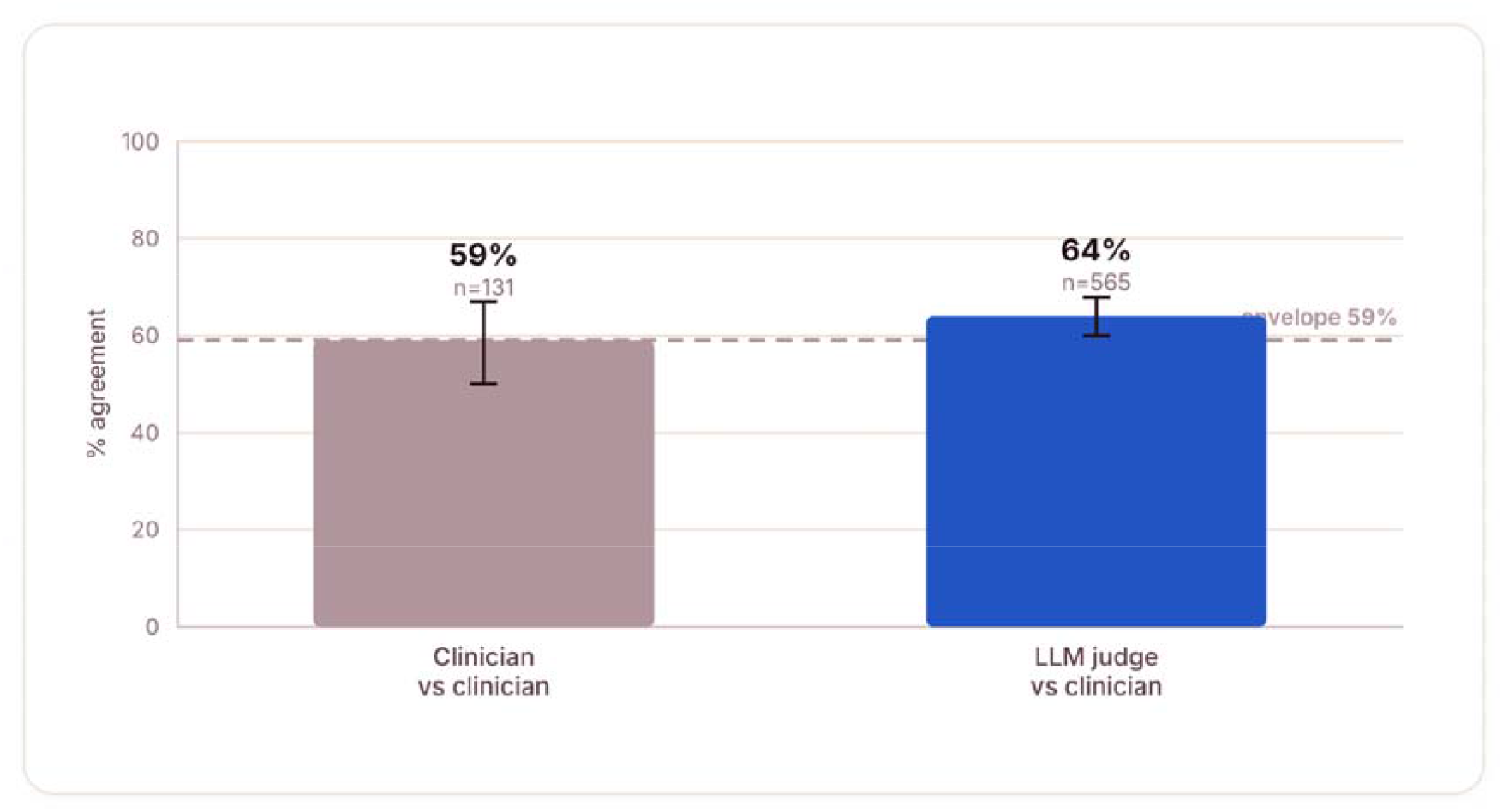
Agreement on genuine error versus not: the judge sits within the inter-clinician envelope. Raw agreement with 95% Wilson intervals. Clinician-clinician agreement (n=131 double-rated) is 59% and judge-clinician agreement (n=565) is 64%; the dashed line marks the inter-clinician envelope.

### Non-differential behaviour across arms

The instrument behaved near-symmetrically on the metrics that determine the parent study’s AI-versus-clinician contrast (figure 2). Human-confirmed kept-precision was 74% for AI-authored items against 81% for clinician-authored items, and the panel-clinician severity signed gap was +0.06 against -0.09 tiers, both small and non-directional. One sub-metric was asymmetric: removed-confirmed, meaning clinician agreement that a screened-out flag was indeed not an error, was 56% for AI items against 42% for clinician items, indicating that the screen over-removes modestly more on clinician-authored notes.

**Figure 2.**
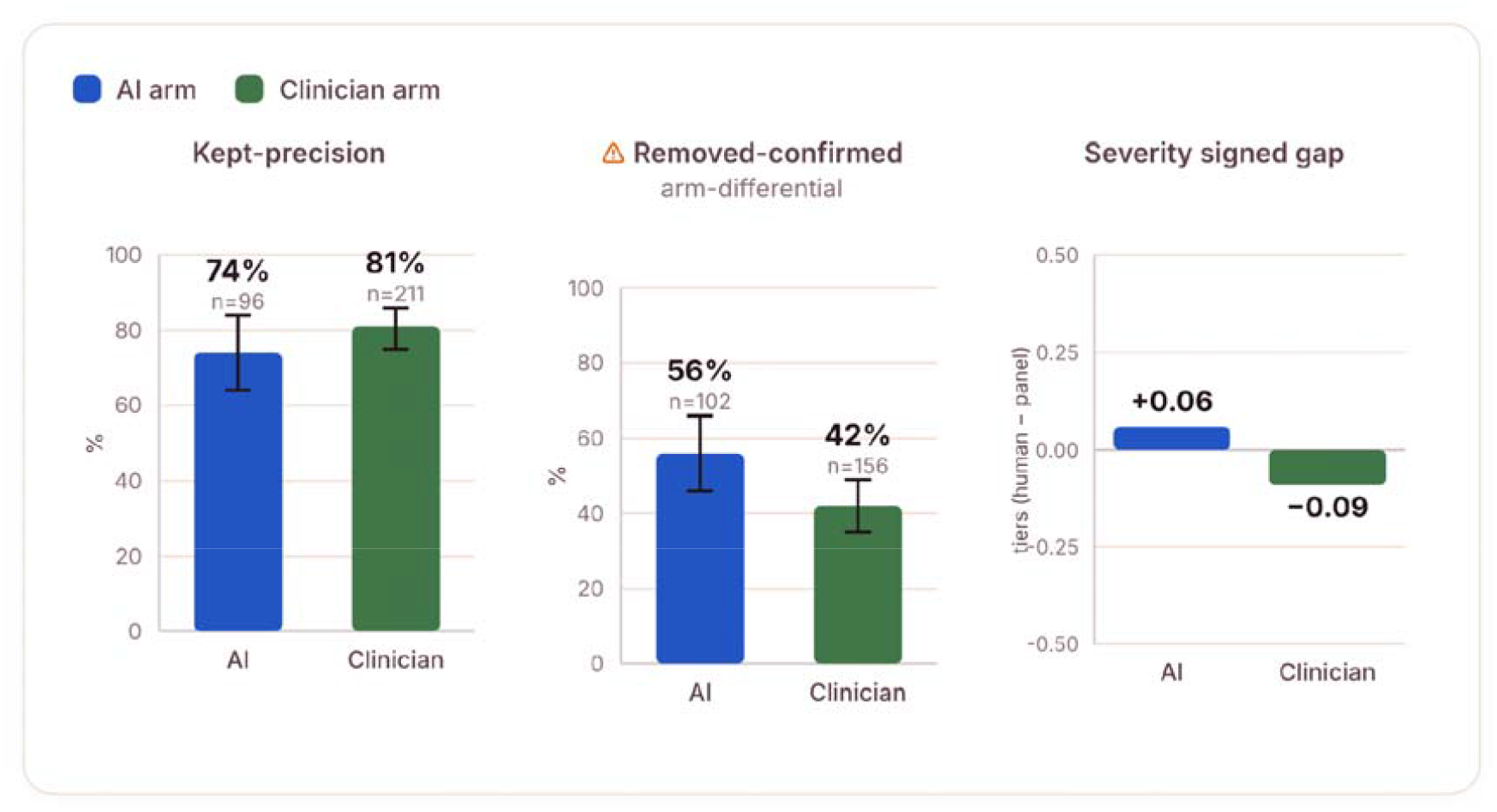
Behaviour across arms: near-symmetric on the contrast-critical metrics, with the false-positive removal rate differing. Three sub-metrics, AI against clinician arm. Kept-precision and the severity signed gap (panel minus clinician, tiers) are near-symmetric. Removed-confirmed is arm-differential: the screen confirms more clinician-note removals as genuine non-errors, meaning it over-removes more on clinician notes. Bars show 95% Wilson intervals where applicable.

The direction of this asymmetry is material and runs against the reported result of the parent study. Because the screen discards proportionally more genuine errors from clinician notes than from AI notes, it understates the clinician-note error rate relative to the AI-note error rate. Any comparative contrast showing clinician notes to carry more errors is therefore conservative with respect to this asymmetry.

### Concordance on clinician-consensus cases

On the 77 items where two clinicians independently agreed, the pipeline concurred on 70% (95% CI 59 to 79), with similar concordance on consensus-genuine items (68%, 95% CI 55 to 79, n=57) and consensus-not-error items (75%, 95% CI 53 to 89, n=20) (figure 3). The judge therefore tracks clinician consensus moderately, consistent with and bounded by the fair inter-clinician agreement reported above, and this does not support a claim of accuracy. The consensus-not-error subset is small and its interval is correspondingly wide.

**Figure 3.**
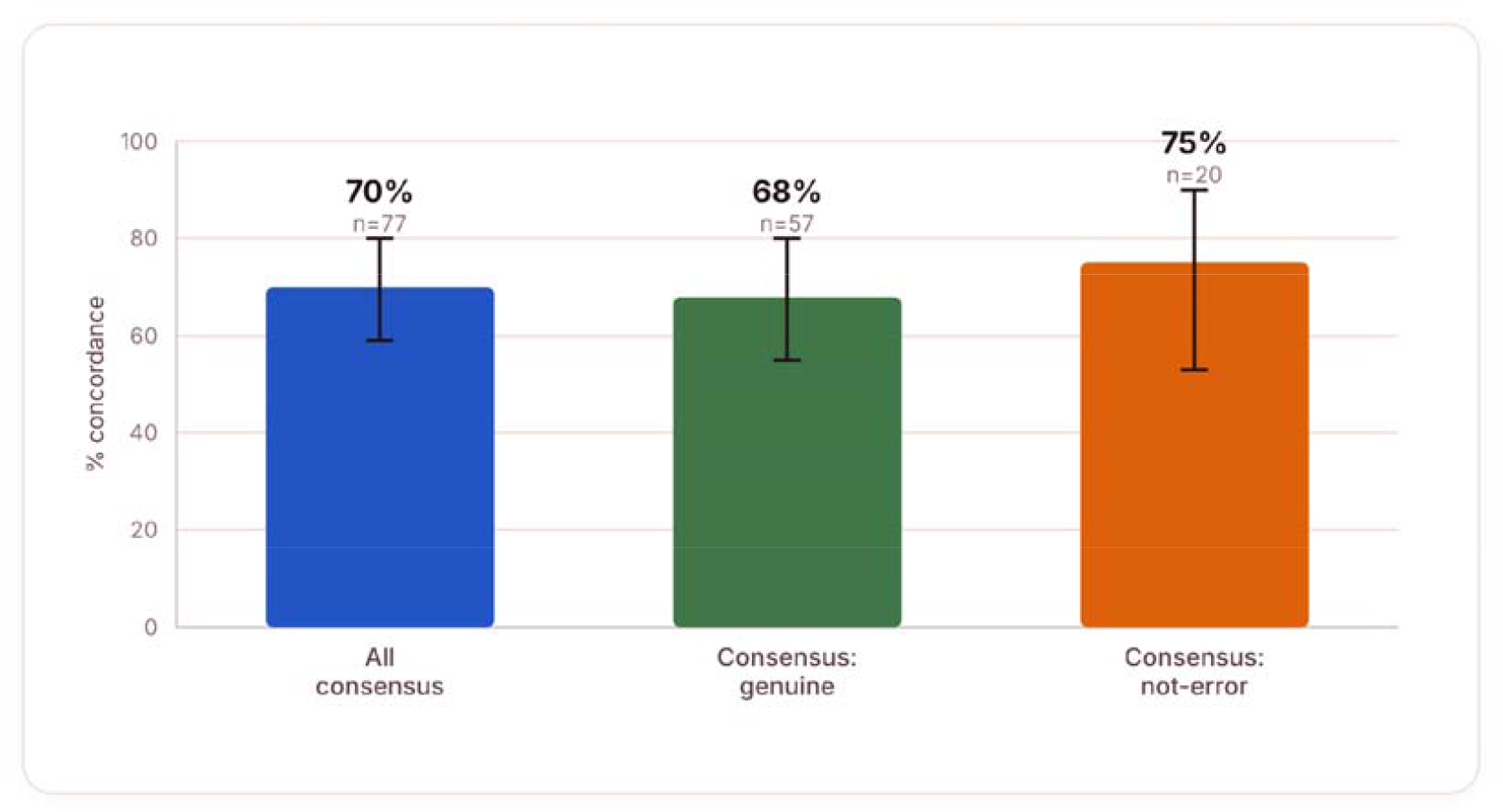
Pipeline concordance where two clinicians agree. Concordance with the clinician-consensus subset, 95% Wilson intervals. Across all consensus items (n=77) the pipeline agrees on 70%, with similar concordance on consensus-genuine (68%, n=57) and consensus-not-error (75%, n=20) cases, bounded by and consistent with the fair inter-clinician agreement.

### Precision recovery and severity grading

Post-screen kept-precision was 79% overall (95% CI 74 to 83, n=307), comprising 77% for fabrication and 80% for omission, which quantifies what the false-positive screen buys. By contrast only 48% of screened-out flags were confirmed as non-errors (95% CI 42 to 54, n=258), which quantifies its cost (supplementary figure S1). Among clinician-confirmed genuine errors, mean Severity x Likelihood was 7.7, placing the average error in the Moderate tier, with 24% reaching the Critical or High tier. The panel and clinician tier distributions were near-identical, with the panel grading marginally more into the Critical and High band than clinicians (28% versus 24%), a conservative direction (figure 4). Clinicians therefore diverge far more on whether an item is an error than on how severe it is once identified.

**Figure 4.**
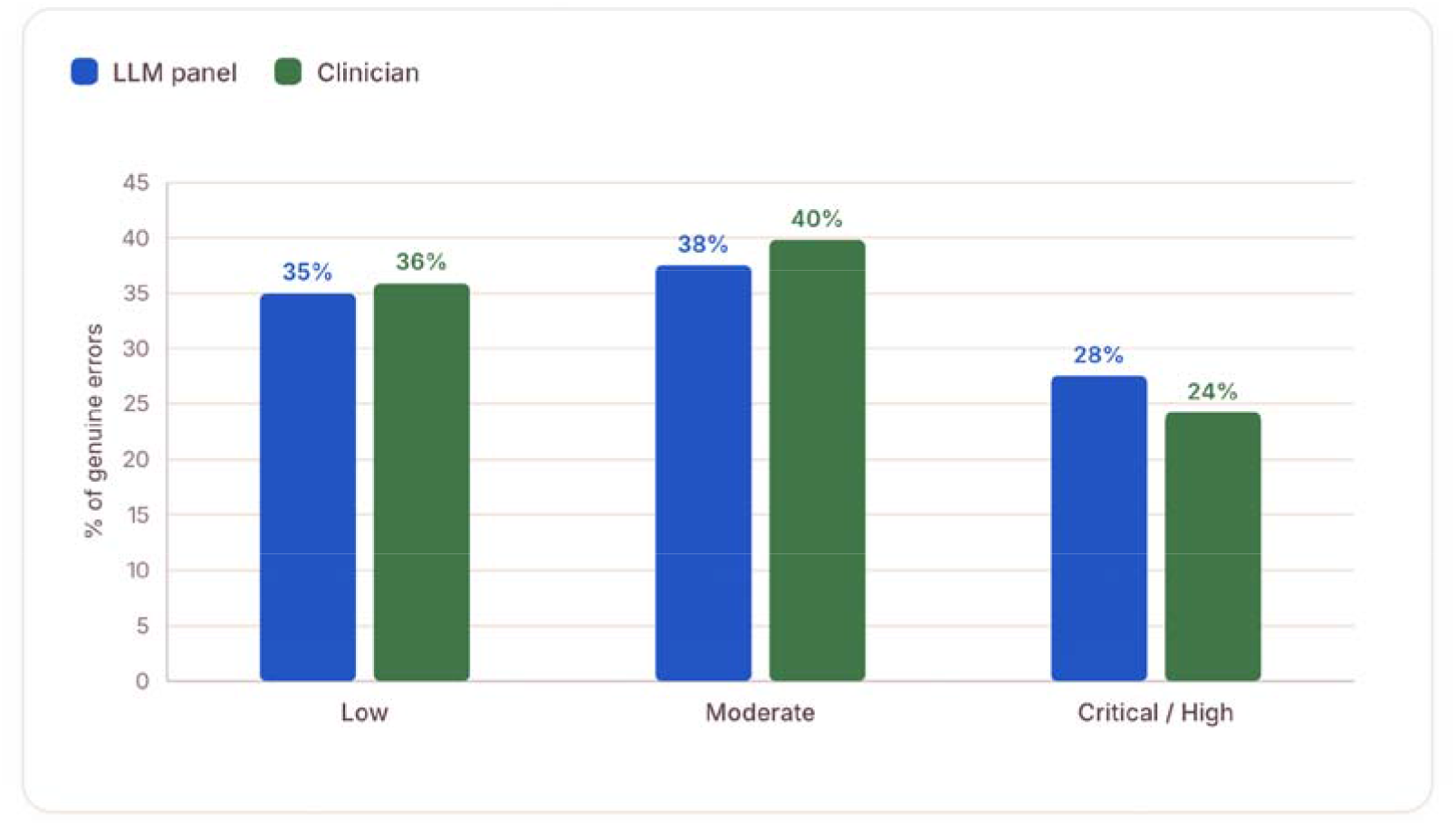
Severity-tier distribution (n=309 genuine and graded): near-identical, so severity grading is non-differential. Distribution of genuine errors across severity tiers, automated panel against clinician. The two distributions are near-identical at Low and Moderate; the panel grades marginally more into the Critical and High band than clinicians (28% against 24%), a conservative direction.

### Robustness to reviewer inclusion

Excluding one reviewer whose independence rests on principal-investigator attestation left the conclusions materially unchanged: kept-precision moved from 79% to 76%, consensus concordance from 70% to 72%, and the non-differential pattern was preserved. Inter-clinician AC1 moved from 0.24 to 0.13, reflecting that this reviewer’s ratings align relatively closely with consensus, which widens the expert envelope and alters no qualitative finding.

### Triangulated genuine-error rate

Because no gold standard exists, we triangulated the genuine-error prevalence among flagged candidates with a Bayesian latent-class model following the Hui-Walter paradigm,^23^ treating the pipeline and clinician calls as two imperfect tests of a latent genuine-error state (supplementary methods; supplementary figures S2 and S3; supplementary table S1). The survey-weighted latent prevalence was 68% (94% credible interval 48 to 83), sitting between the screen’s kept rate (66%) and the clinician genuine rate (74%), and robust to prior specification. This i s a triangulation within the flagged pool, and it does not estimate a population error rate.

### Verdict against the pre-specified decision rule

**Table 1.**
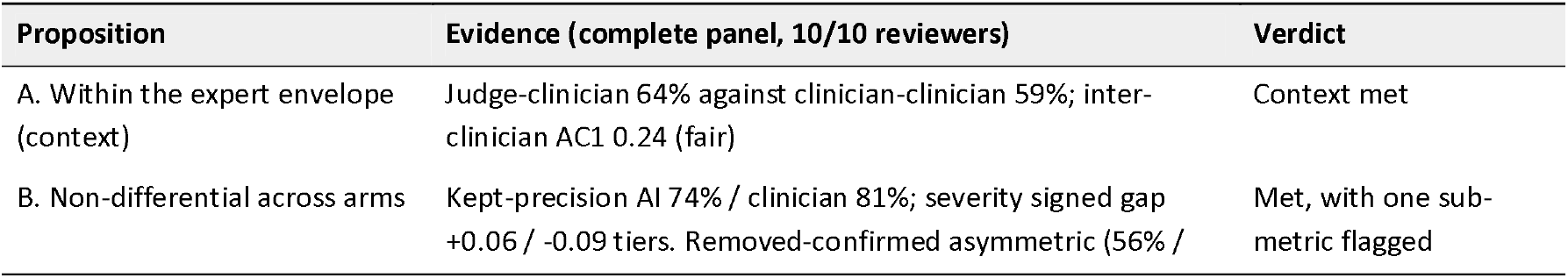
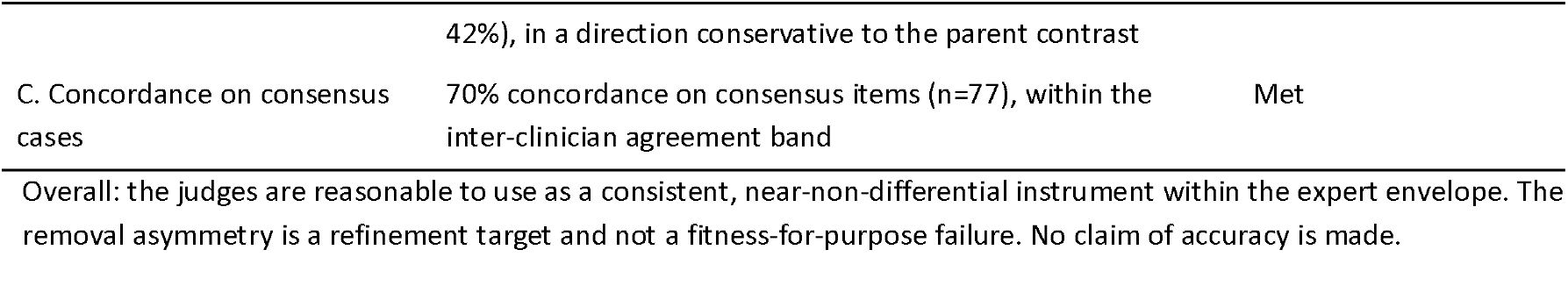
Pre-specified propositions, evidence and verdict.

| Proposition | Evidence (complete panel, 10/10 reviewers) | Verdict |
| --- | --- | --- |
| A. Within the expert envelope (context) | Judge-clinician 64% against clinician-clinician 59%; inter-clinician AC1 0.24 (fair) | Context met |
| B. Non-differential across arms | Kept-precision AI 74% / clinician 81%; severity signed gap +0.06 / -0.09 tiers. Removed-confirmed asymmetric (56% / | Met, with one sub-metric flagged |
|  | 42%), in a direction conservative to the parent contrast |  |
| C. Concordance on consensus cases | 70% concordance on consensus items (n=77), within the inter-clinician agreement band | Met |

Overall: the judges are reasonable to use as a consistent, near-non-differential instrument within the expert envelope. The removal asymmetry is a refinement target and not a fitness-for-purpose failure. No claim of accuracy is made.

## Discussion

### Principal findings

The judges are a consistent, clinician-equivalent instrument that behaves near-symmetrically across the arms it is used to compare, and this licenses a directional AI-versus-clinician comparison. It remains consistent within the expert envelope, which is a weaker property than accuracy.

The licensing argument is a non-differential misclassification argument, and its conditions should be stated explicitly. Where a blinded classifier applies one criterion identically to both arms, and its misclassification is non-differential with respect to arm, the resulting measurement error attenuates the observed contrast toward the null.^24^ That attenuation is not universal: it holds for a dichotomous comparison when misclassification is independent between arms and the classifier performs better than chance, and can exceed the null in some configurations.^24^ Our design satisfies the blinding and identical-criterion conditions, and the data support approximate non-differentiality on kept-precision and severity. A contrast surviving such an instrument is therefore conservative under those conditions, which we state explicitly, since attenuation should not be treated as automatic.

### The honest ceiling

Consistency and within-envelope parity are not accurate. Moderate consensus-case concordance (70%) and fair inter-rater reliability (AC1 0.24) preclude any claim that the judge is correct. The fair inter-clinician agreement is itself the most consequential finding, because it reframes how scribe-error instruments should be validated: against the envelope of expert judgement, since no truth exists to validate against.

### Comparison with other studies

Our agreement figures are substantially lower than those reported for large language model judges of clinical text quality, where an intraclass correlation of 0.818 (95% CI 0.772 to 0.854) against human evaluators has been achieved on a validated ordinal summarisation-quality instrument.^9^ We do not read this as a contradiction. Those studies ask raters to score the overall quality of a document on an ordinal scale, a global judgement on which raters converge readily; we ask whether one specific span of text constitutes a genuine documentation error, an item-level binary judgement requiring a rater to place the boundary between a faithful paraphrase and an omission. The case-note audit literature predicts this gap, having found reliability higher for explicit criteria and outcome judgements and lower for implicit criteria and process errors^6^. Error detection therefore appears to be the harder measurement problem for this class of instrument, and reliability figures from the two tasks should not be pooled. Our finding that clinicians converge on severity while diverging on genuineness supports the same reading.

The direction of our severity findings is also consistent with the wider literature: comparative studies of generated clinical documentation report fabrication and omission in clinician-authored as well as AI-authored material,^3 25^ which is the pattern an instrument that is non-differential across arms would be expected to reproduce.

### Bounded truth

The genuine-error rate should be reported as an interval, with the false-positive-stripped human-identification arm as a lower bound and the precision-adjusted automated arm as an upper bound, and the location of the truth between them explicitly left unestimated. The latent-class triangulation places the rate among flagged candidates at 68% (94% credible interval 48 to 83), between the screen (66%) and clinician (74%) rates, though this is a within-pool triangulation and not a population error rate.

### Limitations

The one asymmetry we observed is the removal differential: the screen over-removes modestly more on clinician-authored notes. This is a content behaviour and not a strictness one, and we treat it as a refinement target for a future instrument version, to be addressed by parent-study re-scoring and a locked-rule re-test. As reported above, its direction is conservative with respect to the parent study’s finding, which is the relevant consideration for whether the instrument may be used, though a reader who wishes to treat proposition B as only partially met is entitled to do so on this evidence.

Precision is limited by the double-rated subsample. Inter-rater reliability rests on 131 items and ten reviewers, so the AC1 point estimate carries appreciable uncertainty.

The binary genuineness decision was forced, with no uncertain option. This was a deliberate design choice to obtain a usable signal, but it converts genuine reviewer uncertainty into apparent disagreement and may therefore depress the observed inter-clinician agreement. The fair AC1 should be read with that in mind, and a graded-confidence elicitation would be a reasonable design variation for future work.

Reviewers were post-certification general practitioners with emergency and surgical representation, adjudicating material drawn from a mixed case-mix. Where an item concerned content outside a reviewer’s routine scope, adjudication may be less reliable, and we did not stratify agreement by reviewer specialty against item content.

The latent-class triangulation assumes conditional independence between the pipeline and clinician calls. This is imperfect given shared model lineage in the pipeline, and where classifier errors are positively correlated given the latent state, latent-class models overstate test accuracy and can bias the prevalence estimate.^22^ The estimate is a triangulation and not a verdict, and its wide credible interval reflects genuine uncertainty from limited double-rating and weak inter-clinician agreement.

Validation was restricted to a single language and setting, so cross-language instrument behaviour is uncharacterised. One reviewer’s independence rests on principal-investigator attestation and is reported with a robustness analysis. Finally, the clinician comparator is itself imperfect by design, so all estimands are characterisations of instrument behaviour or bounds, and never accuracy.

## Conclusion

Where no gold standard can exist, the right test of an automated documentation-error judge is not accuracy but defensibility: reproducibility, operation within the envelope of expert disagreement, and non-differential behaviour across the arms it is used to compare. In this pre-registered, blinded human-validation study, the false-positive screen and severity panel meet that bar. They agree with clinicians as well as clinicians agree with one another, behave near-symmetrically on the contrast-critical metrics, and track clinician consensus moderately. The instrument is therefore reasonable to use to measure a comparative AI-versus-clinician contrast, with its absolute error rate reported as an interval and its one removal asymmetry, whose direction is conservative, flagged for refinement.

## Contributors

HB conceived and designed the study, developed the methodology and analysis plan, conducted the analysis, and drafted the manuscript. VL contributed to the methodology and validation, critically revised the manuscript, and contributed to its review and editing. BA contributed to the methodology, regulatory framing, validation, and risk-grading framework, and critically revised the manuscript. RS developed and implemented the software and analysis pipeline, contributed to the methodology, investigation, data curation, validation and visualisation, and reviewed and edited the manuscript. All authors approved the final version and accepted responsibility for the decision to submit. HB is the guarantor.

## Funding

This study was funded in its entirety by Heidi Health. No external, government or third-party funding was received. All authors are employees of the funder, which was therefore involved in the study design, the conduct of the study, the analysis and interpretation of the data, the preparation of the manuscript and the decision to submit it for publication.

## Competing interests

All authors are employees of Heidi Health, which develops the ambient AI scribe and automated judges evaluated in this study. The ten adjudicating clinicians were independent of Heidi Health and blinded to note authorship and the automated pipeline’s verdict. The analysis plan and decision rule were fixed before data collection.

## Patient consent for publication

Not required. No patients were involved and no patient-identifiable data were used.

## Ethics and consent

This study involved no patients and no patient data. It re-analysed candidate error records generated by a simulation study using scripted consultations performed by trained actors. Adjudicating clinicians participated as professional reviewers and provided informed consent to the research use of their judgements.

## Data availability statement

Data supporting the findings of this study are reported in the article and accompanying Supplementary Materials. Additional study materials are provided where appropriate to support interpretation and reproducibility. The underlying evaluation datasets, source materials, and analysis or evaluation code may contain proprietary or commercially sensitive information and are not publicly available. Enquiries regarding access to additional materials may be directed to the corresponding author and will be considered subject to applicable confidentiality, intellectual-property, data-governance, and commercial requirements.

## Use of AI-assisted technologies

The manuscript was drafted and substantively edited by the named authors. No external writing assistance was provided or funded. Artificial intelligence and large language models were evaluated as part of the study methodology; however, no generative AI, machine learning, or large language model tools were used in the authorship, drafting, or substantive editing of the manuscript.

## Notes

### Author Declarations

No patients or patient data were involved. Source materials comprised candidate error records generated from scripted simulated consultations. The ten independent external clinicians participated as professional adjudicators and provided informed consent for the research use of their judgements.

