## Supplemental File for "Are automated documentation-error judges fit to measure ambient AI scribes? A pre-registered, blinded human-validation study"

Bergman H, Liu V, Austin B, Sanghera R

References cited here are numbered independently of the main text and listed in full at the end of this document.

**Contents**

| **Item** | **Title** | **Cited in main text at** |
| --- | --- | --- |
| Supplementary methods | Bayesian latent-class triangulation of the genuine-error rate: rationale, model, estimation and assumptions | Results, triangulated genuine-error rate |
| Table S1 | Latent-class posterior estimates for instrument operating characteristics and genuine-error prevalence | Results, triangulated genuine-error rate |
| Figure S1 | Precision recovery: human-confirmed rates for retained and removed flags | Results, precision recovery and severity grading |
| Figure S2 | Triangulating the genuine-error rate within the flagged pool | Results, triangulated genuine-error rate |
| Figure S3 | Estimated instrument operating characteristics from the latent-class model | Results, triangulated genuine-error rate |

**Supplementary methods. Bayesian latent-class triangulation**

**Rationale**

The main analysis establishes that neither the automated pipeline nor expert clinicians constitute a gold standard for whether a flagged item is a genuine documentation error. Where no reference is available, the accuracy of multiple imperfect classifiers and the latent prevalence they are jointly measuring can be estimated by a Bayesian latent-class model following the Hui-Walter paradigm.¹ We use it as a triangulation to place a caveated value on the probable middle between the two instruments, and not as a definitive estimate.

**Model**

For each flagged candidate we observe two imperfect tests of a latent binary state, namely whether the item is a genuine documentation error: the pipeline, where a retained flag is a positive and a removed flag a negative, and one or two clinician adjudications. Conditional on the latent state, the pipeline and clinician calls are assumed independent, and the two clinician reads are assumed independent and exchangeable with shared sensitivity and specificity. The latent class is marginalised analytically. The parameters are the prevalence of genuine errors, the pipeline's sensitivity and specificity, and a clinician's sensitivity and specificity.

The double-rated items (n=131) provide the clinician-by-clinician information that identifies the clinician parameters; single-rated items contribute the pipeline-by-clinician cross-tabulation. The model was fitted in PyMC using the No-U-Turn Sampler with two chains of 1,500 draws each, and weakly informative Beta priors encoding that each test performs better than chance, which resolves latent-label switching. Because the sample is stratified, the latent prevalence is projected to the full flagged pool by survey weighting each item by the ratio of natural to sampled stratum size, applied to its posterior class-membership probability.

**Table S1. Latent-class posterior estimates**

| **Parameter** | **Posterior mean** | **94% credible interval** |
| --- | --- | --- |
| Pipeline sensitivity | 0.83 | 0.68 to 0.98 |
| Pipeline specificity | 0.87 | 0.68 to 0.99 |
| Clinician sensitivity | 0.82 | 0.75 to 0.91 |
| Clinician specificity | 0.57 | 0.45 to 0.75 |
| Genuine-error prevalence (survey-weighted, flagged pool) | 0.68 | 0.48 to 0.83 |

Complete panel; PyMC, No-U-Turn Sampler, weakly informative priors. Observed survey-weighted positive rates were 66% for the pipeline screen and 74% for clinicians. The latent estimate of 68% sits between them and was robust to a more diffuse prior set (0.67, 94% credible interval 0.48 to 0.83). The clinician specificity estimate of 0.57 is the lowest operating characteristic in the model, which is consistent with clinicians confirming fewer screen-removed flags as genuine non-errors.

**Figure S1. Precision recovery**

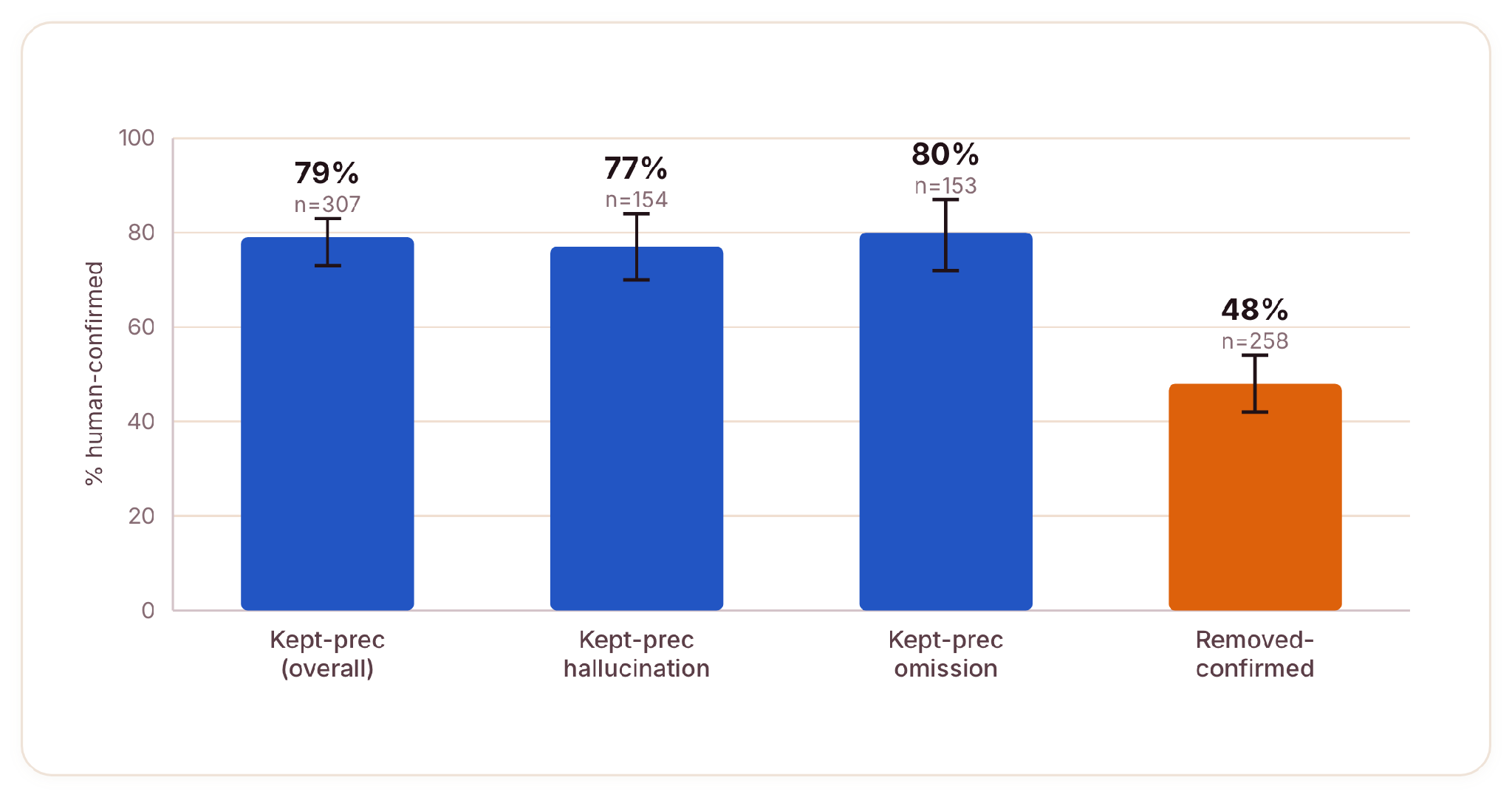

**Figure S1 Human-confirmed rates for retained and removed flags, with 95% Wilson intervals.**

Retained flags are confirmed genuine at 79% overall and similarly across fabrication (77%) and omission (80%), which is the precision the false-positive screen recovers. By contrast only 48% of screen-removed flags are confirmed as non-errors (n=258), which quantifies the screen's cost.

**Figure S2. Triangulating the genuine-error rate**

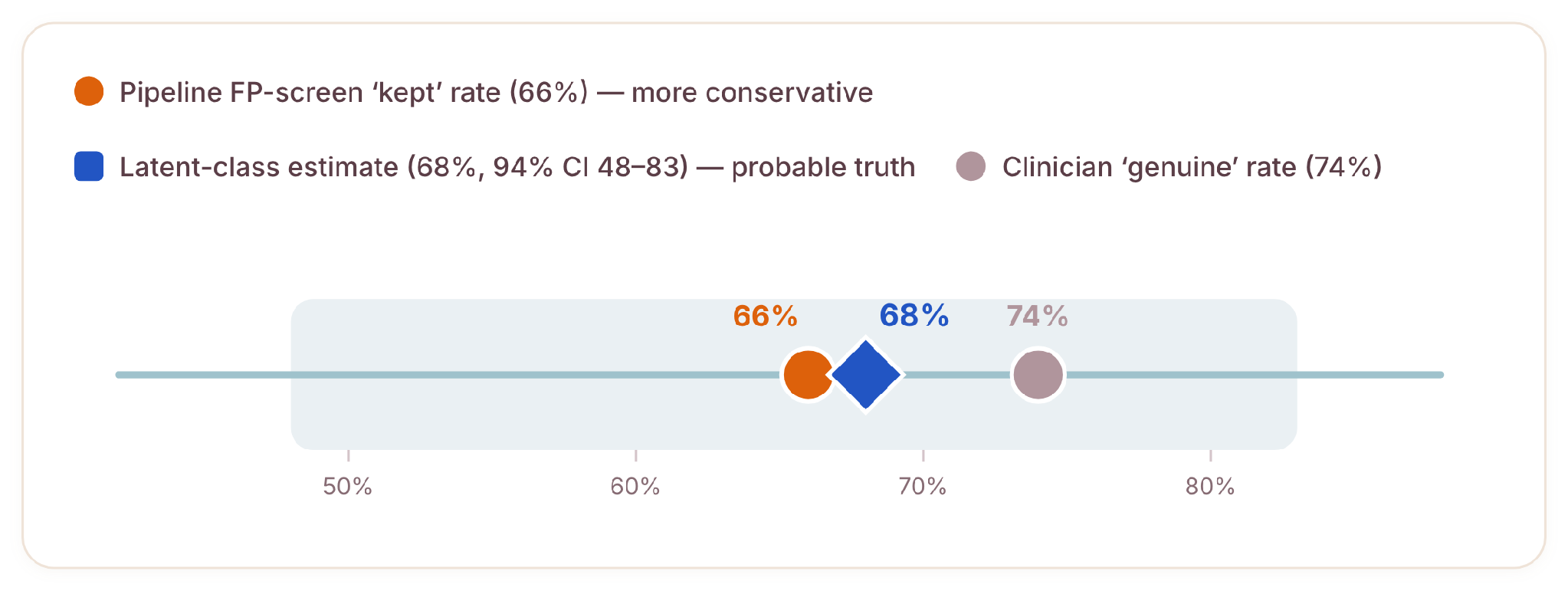

**Figure S2 The latent estimate (68%, 94% credible interval 48 to 83) sits between the screen and clinician rates.**

Latent-class, survey-weighted; prevalence among flagged candidates only and not a population rate. The shaded band is the 94% credible interval. The two instruments bracket the latent estimate closely. The intuitive contrast between an over-sensitive automated judge and an under-sensitive clinician operates at the identification stage, which lies outside this flagged pool.

**Figure S3. Estimated instrument operating characteristics**

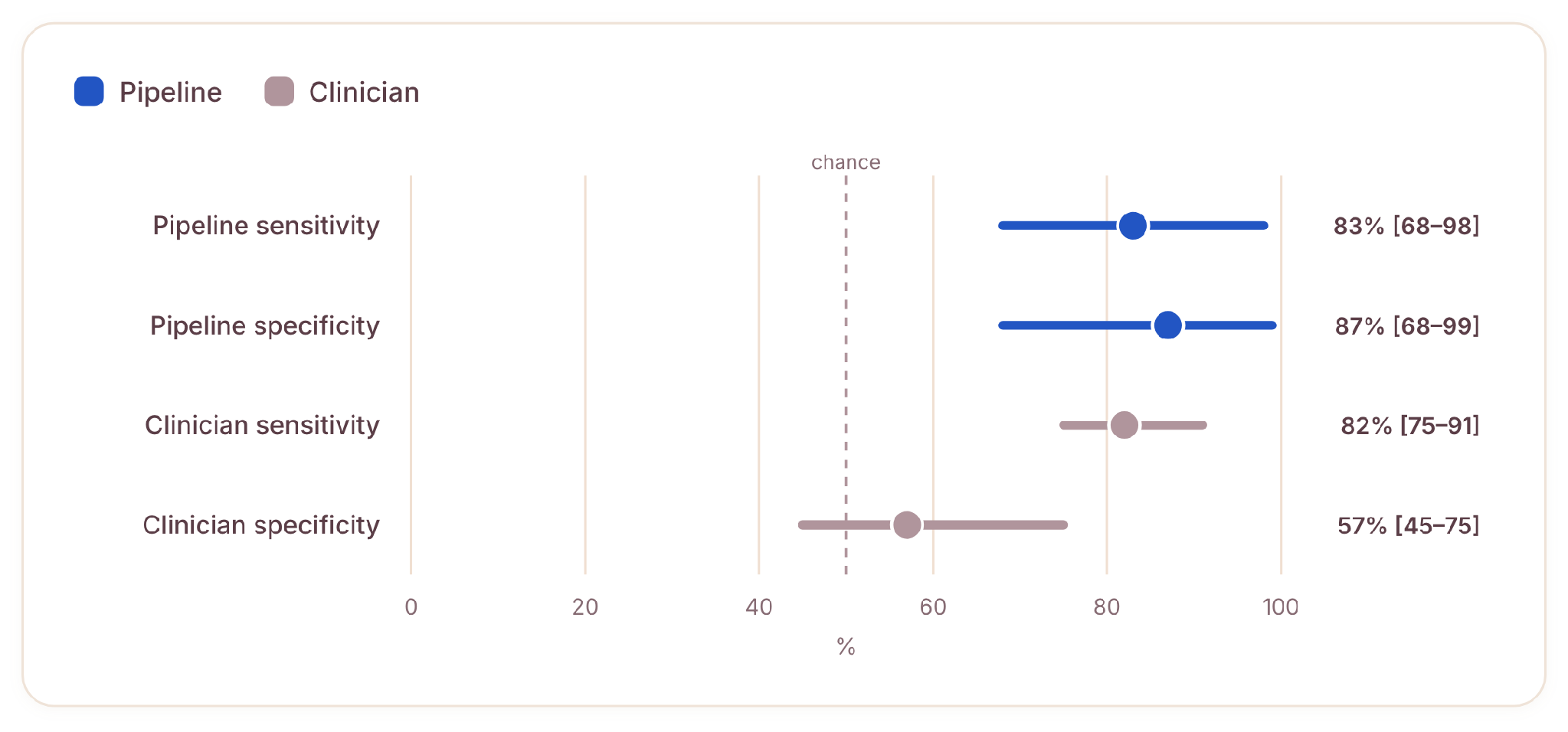

**Figure S3 Posterior means with 94% credible intervals; the dashed line marks chance.**

Wide intervals reflect genuine uncertainty arising from limited double-rating and weak inter-clinician agreement, so these are triangulated estimates and not a verdict on either instrument. The clinician specificity estimate is the lowest operating characteristic in the model.

**Assumptions and limitations of the triangulation**

**Prevalence among flagged candidates, not the population error rate.** The sample is conditioned on the pipeline having flagged the item, so there is no cell for items missed by both instruments and errors absent from the candidate pool are not estimable. This estimate must not be read as a rate of documentation errors per note.

**Conditional independence is imperfect.** The false-positive screen and the automated identifier share model lineage. Where classifier errors are positively correlated given the latent state, latent-class models overstate test accuracy and can bias the prevalence estimate,² so the estimate is a triangulation and not a verdict.

**Wide credible intervals.** The prevalence interval of 0.48 to 0.83 reflects genuine uncertainty from limited double-rating and weak inter-clinician agreement.

**Approximate survey weights.** Weights were computed at the level of the decision-by-tier-by-source-by-type stratum.

**Reproducibility**

Analysis seed 20260518. Inputs are the adjudication responses table and the item-level key; outputs are Figures S2 and S3 and the results file. Complete panel: 565 adjudications from 10 reviewers across 434 flagged items, 131 of them double-rated.
